# High Consistency, Limited Accuracy: Evaluating Large Language Models for Binary Medical Diagnosis

**DOI:** 10.64898/2025.12.08.25341823

**Authors:** Dwi Anggriani, Syaiful Bachri Mustamin, Muhammad Atnang, Kartini Aprilia Pratiwi Nuzry

**Author notes:** **Corresponding Author:** Syaiful Bachri Mustamin.

## Abstract

**Background:** Large Language Models (LLMs) have demonstrated impressive capabilities in medical knowledge tasks, achieving 60-80% accuracy on licensing examinations. However, their reliability and consistency in clinical diagnosis—critical for clinical trustworthiness–remain incompletely characterized.

**Objective:** To systematically evaluate the consistency and diagnostic accuracy of state-of-the-art LLMs in binary medical diagnosis, examining the relationship between reproducibility and diagnostic performance.

**Methods:** We evaluated three frontier LLMs (GPT-40, Gemini-2.0-Flash, Qwen-Plus) on heart disease diagnosis using 100 diverse clinical cases from the UCI Heart Disease dataset. Each model performed 4 independent assessments per case (1,200 total predictions). We tested two prompt variations (“Expert Cardiologist” vs “Neutral Assessor”) and measured intra-model consistency, inter-model agreement, diagnostic accuracy, and prompt sensitivity using a SQLite-based checkpoint system.

**Results:** All models achieved exceptional intra-model consistency (99-100%), with Qwen-Plus demonstrating perfect reproducibility (100%). Inter-model agreement was similarly high (98-99%), indicating convergent reasoning patterns. However, diagnostic accuracy remained at approximately 50%, equivalent to random guessing. Models exhibited strong systematic bias toward positive diagnosis (49-51 false positives vs 0-1 false negatives per 100 cases). Prompt variation had minimal impact (≤3% prediction changes), and error patterns were highly systematic, with all models making identical errors on 48-51% of cases. This created a consistency-accuracy gap of approximately 50 percentage points.

**Conclusions:** Our findings reveal a critical dissociation between consistency and accuracy in LLM medical diagnosis. While LLMs demonstrate remarkable reproducibility–a desirable property for clinical tools–their systematic tendency toward over-diagnosis and limited discriminative accuracy constrain direct clinical utility. The high inter-model agreement on errors suggests fundamental limitations in applying general-purpose LLMs to medical diagnosis rather than model-specific artifacts. Results suggest LLMs may be better suited as supplementary decision-support tools with human oversight rather than primary diagnostic systems. Future development should prioritize discriminative fine-tuning on labeled diagnostic datasets and calibration techniques to address systematic biases.

## INTRODUCTION

Large Language Models (LLMs) have emerged as promising tools for clinical applications, demonstrating impressive performance on medical licensing examinations and case analysis [1-3]. Recent evaluations show frontier models achieving 60-80% accuracy on USMLE exams and other standardized medical assessments, approaching or exceeding average human physician performance on knowledge-based tasks. However, their deployment in clinical settings raises critical questions about reliability and consistency.

Unlike traditional diagnostic tools expected to yield reproducible results, LLMs employ stochastic generation mechanisms that can lead to varying outputs for identical inputs. Temperature parameters, sampling methods, and prompt formulations can all influence model behavior, potentially undermining clinical reproducibility. Despite this concern, most evaluations of LLMs in medical contexts focus on single-run accuracy without assessing reproducibility across repeated assessments [4].

### Research Gap

The existing literature demonstrates a significant gap in understanding the relationship between consistency and accuracy in LLM medical diagnosis. While studies have evaluated diagnostic accuracy on various medical tasks [5,6], few have systematically examined whether high accuracy is accompanied by high consistency, or whether models can be consistently wrong. Additionally, the influence of prompt engineering-often cited as a method to improve LLM performance-remainsincompletely characterized for diagnostic tasks [7].

Three key questions remain unanswered:

1. **How reproducible are LLM diagnostic assessments?** Do models provide consistent diagnoses when presented with identical clinical information multiple times?
2. **What is the relationship between consistency and accuracy?** Can models be highly consistent but systematically inaccurate, or does consistency guarantee accuracy?
3. **How sensitive are diagnostic decisions to prompt formulation?** Can prompt engineering effectively modulate diagnostic behavior, or is behavior deeply encoded in model weights?

### Study Objectives

This study addresses these gaps through comprehensive evaluation of three state of-the-art LLMs on binary heart disease diagnosis. Our specific aims were to:

1. **Quantify intra-model consistency** by measuring agreement across repeated independent assessments of identical cases
2. **Evaluate inter-model agreement** to determine whether diagnostic patterns are model-specific or generalizable
3. **Measure diagnostic accuracy** relative to ground truth and compare with consistency metrics
4. **Assess prompt sensitivity** by comparing two distinct prompt formulations
5. **Analyze error patterns** to determine whether mistakes are random or systematic

Understanding these relationships is critical for responsible deployment of LLMs in clinical settings, where both accuracy and reproducibility are essential for patient safety and physician trust.

## METHODS

### Dataset and Study Design

We utilized the UCI Heart Disease dataset [8], a widely-used benchmark containing clinical data from 303 patients evaluated for coronary artery disease. The dataset includes 13 clinical parameters:

- **Demographics:** Age, sex
- **Symptoms:** Chest pain type (4 categories: typical angina, atypical angina, non-anginal pain, asymptomatic)
- **Vital signs:** Resting blood pressure (mm Hg)
- **Laboratory values:** Serum cholesterol (mg/dl), fasting blood sugar (>120 mg/dl: yes/no)
- **Electrocardiography:** Resting ECG findings (normal, ST-T wave abnormality, left ventricular hypertrophy)
- **Exercise testing:** Maximum heart rate achieved (beats/min), exercise induced angina (yes/no), ST depression induced by exercise relative to rest (mm), slope of peak exercise ST segment (upsloping, flat, downsloping)
- **Imaging:** Number of major vessels (0-3) colored byfluoroscopy
- **Thalassemia test:** Normal, fixed defect, reversible defect

The binary outcome indicated presence (1) or absence (o) of significant coronary artery stenosis based on angiography.

To ensure diverse representation across the clinical spectrum, we performed k means clustering (k=2) on all features and selected 50 cases from each cluster via stratified random sampling. This yielded 100 test cases with balanced disease prevalence (51% positive, 49% negative), representing varied clinical presentations from low-risk to high-risk profiles.

### Models Evaluated

We evaluated three frontier LLMs representing different architectural families and training approaches:

1. **GPT-40** (OpenAI): Multimodal model with extensive medical training
2. **Gemini-2.0-Flash**(Google): Efficient variant with strong reasoning capabilities
3. **Qwen-Plus** (Alibaba): Large-scale Chinese-English bilingual model

All models were accessed via official APis with temperature=o.7 to balance determinism and natural language generation. Each API call was independent, ensuring no information leakage between runs.

### Prompt Design

We tested two prompt formulations to assess sensitivity to framing:

### Prompt A (“Expert Cardiologist”)

~~~
You are Dr. CardioExpert, a highly experienced cardiologist with over
20 years of
clinical practice. You are reviewing a patient’s clinical data to determine if they
have heart disease. Based on the following clinical parameters, provide your diagnosis.
[Clinical data provided]
Does this patient have heart disease? Answer with:
“Yes” if you believe heart disease is present
“No” if you believe heart disease is absent
Provide a brief 2-3 sentence justification for your diagnosis.
~~~

### Prompt B (“Neutral Assessor”)

~~~
You are a medical AI assistant trained to provide accurate and
balanced diagnostic
assessments. Your goal is to carefully evaluate clinical data and
provide a diagnosis,
avoiding both over-diagnosis and under-diagnosis. Based on the
following clinical
parameters, provide your diagnosis.
[Clinical data provided]
Does this patient have heart disease? Answer with:
“Yes” if the clinical evidence suggests heart disease is present
“No” if the clinical evidence suggests heart disease is absent
Provide a brief 2-3 sentence justification for your diagnosis.
~~~

Both prompts provided identical clinical data with clear parameter definitions. The key difference was framing: Prompt A emphasized expert authority (potentially inducing medical conservatism), while Prompt B emphasized balanced assessment.

### Experimental Protocol

Each model performed 4 independent diagnostic assessments for each of the 100 test cases, yielding 1,200 predictions per prompt (100 cases × 4 runs × 3 models). We implemented a SQLite-based checkpoint system with the following features:

- **Immediate data persistence:** Each prediction saved immediately after API response
- **Duplicate prevention:** UNIQUE constraint on (test_id, run_id, model) prevented accidental re-runs
- **Automatic resumption:** System detected completed runs and continued from last checkpoint
- **Comprehensive logging:** Timestamps, justifications, and error messages recorded for all predictions

The system enabled reliable execution across multiple days despite API rate limits and connection issues, ensuring complete data collection.

### Outcome Measures

#### Primary outcomes

1. **Intra-model consistency:** For each case, we calculated the proportion of 4 runs with majority agreement (≥2/4 identical predictions). Perfect consistency = 100% (all 4 runs identical).
2. **Diagnostic accuracy:** Using majority voting per case ( ≥2/4runs for final diagnosis), we calculated:
  - Accuracy: (TP +TN)/ Total
  - Sensitivity (Recall): TP / (TP + FN)
  - Specificity: TN / (TN + FP)
  - Precision: TP / (TP + FP)
  - Fi-score: 2 × (Precision × Recall) / (Precision + Recall)
3. **Inter-model agreement:** Pairwise agreement between models and Cohen’s kappa for chance-corrected agreement

#### Secondary outcomes

**4. Prompt sensitivity:** Proportion of cases with identical predictions across both prompt formulations

**5. Error pattern analysis:** Classification of cases as all-correct (all 3 models right), all-wrong (all 3 models wrong), or mixed outcomes

### Statistical Analysis

We calculated descriptive statistics (mean, standard deviation, range) for all metrics. Confusion matrices visualized diagnostic patterns. Cohen’s kappa assessed inter-model agreement with chance correction. All analyses used Python with pandas, NumPy, scikit-learn, and scipy. Statistical significance was set at p<0.05 (two-tailed tests).

## RESULTS

### 1. Intra-Model Consistency: Exceptional Reproducibility

All three models demonstrated remarkably high intra-model consistency across repeated assessments (Table 1). Qwen-Plus achieved perfect consistency (100%) with Prompt A, never varying across 4 independent runs for any of the 100 cases. GPT-40 and Gemini-2.0-Flash showed 99.0-99.5% average consistency.

**Table 1.** Intra-Model Consistency Metrics.

| Model | Prompt | Avg<br>Consistency<br>(%) | Min<br>Consistency<br>(%) | Perfect<br>Agreement<br>(%) |
| --- | --- | --- | --- | --- |
| GPT-4o | Expert | 99.25 | 50.0 | 98 |
| GPT-4o | Neutral | 99.00 | 75.0 | 96 |
| Gemini-2.0-Flash | Expert | 99.50 | 75.0 | 98 |
| Gemini-2.0-Flash | Neutral | 99.25 | 50.0 | 98 |
| Qwen-Plus | Expert | **100.00** | 100.0 | 100 |
| Qwen-Plus | Neutral | 99.75 | 75.0 | 99 |

*Perfect Agreement: Proportion of cases with all 4 runs identical*

Notably, 96-100% of cases achieved perfect agreement (4/ 4 identical predictions), and minimum consistency never fell below 50% (indicating at least 2/4 runs agreed in all cases). This demonstrates that LLMs apply reasoning patterns consistently rather than generating random outputs.

### 2. Inter-Model Agreement: High Consensus

Models showed 98-100% pairwise agreement, indicating remarkably similar reasoning patterns despite different architectures and training procedures (Table 2).

**Table 2.** Inter-Model Agreement.

| Prompt | GPT-Gemini | GPT-Qwen | Gemini-Qwen | 3-Way<br>Agreement<br>(%) | Cohen's<br>$\kappa$ |
| --- | --- | --- | --- | --- | --- |
| Expert | 98.0% | 100.0% | 98.0% | 98 | 0.01 |
| Neutral | 100.0% | 99.0% | 99.0% | 99 | 0.03 |

*3-Way Agreement: Proportion of cases where all 3 models provided identical diagnosis*

Three-way agreement (all models concurring) occurred in 98-99% of cases. Cohen’s kappa values near zero reflected extreme class imbalance (nearly all positive predictions) rather than lack of agreement-when one model predicted “disease,” others almost always agreed.

### 3. Diagnostic Accuracy: Limited Despite High Consistency

Diagnostic accuracy approximated random guessing (48-51%) despite 99-100% consistency (Table 3). This created a consistency-accuracy gap of approximately 50 percentage points.

**Table 3.**
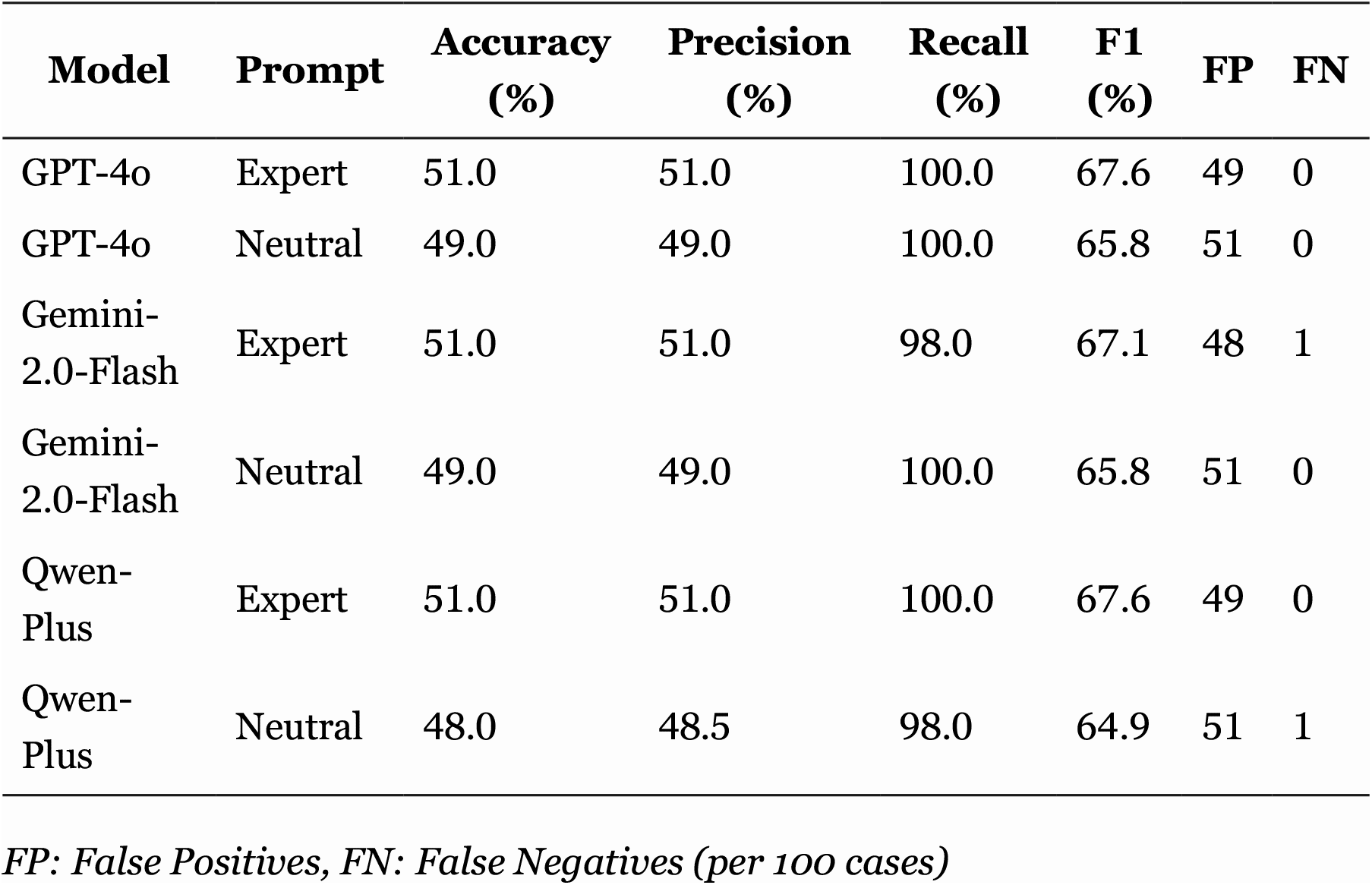
Diagnostic Performance Metrics.

| Model | Prompt | Accuracy (%) | Precision (%) | Recall (%) | F1 (%) | FP | FN |
| --- | --- | --- | --- | --- | --- | --- | --- |
| GPT-4o | Expert | 51.0 | 51.0 | 100.0 | 67.6 | 49 | 0 |
| GPT-4o | Neutral | 49.0 | 49.0 | 100.0 | 65.8 | 51 | 0 |
| Gemini-2.0-Flash | Expert | 51.0 | 51.0 | 98.0 | 67.1 | 48 | 1 |
| Gemini-2.0-Flash | Neutral | 49.0 | 49.0 | 100.0 | 65.8 | 51 | 0 |
| Qwen-Plus | Expert | 51.0 | 51.0 | 100.0 | 67.6 | 49 | 0 |
| Qwen-Plus | Neutral | 48.0 | 48.5 | 98.0 | 64.9 | 51 | 1 |

*FP: False Positives, FN: False Negatives (per 100 cases)*

Models achieved perfect or near-perfect recall (98-100%), indicating excellent sensitivity for detecting disease. However, specificity was extremely poor (∼o-2%), generating 49-51false positives versus only 0-1 false negatives. This pattern suggests systematic positive diagnosis bias rather than random errors.

Representative confusion matrices (Figure 1) showed models predicted “disease present” for nearly all cases, with true negatives ≈o across all model-prompt combinations.

**Figure 1.**
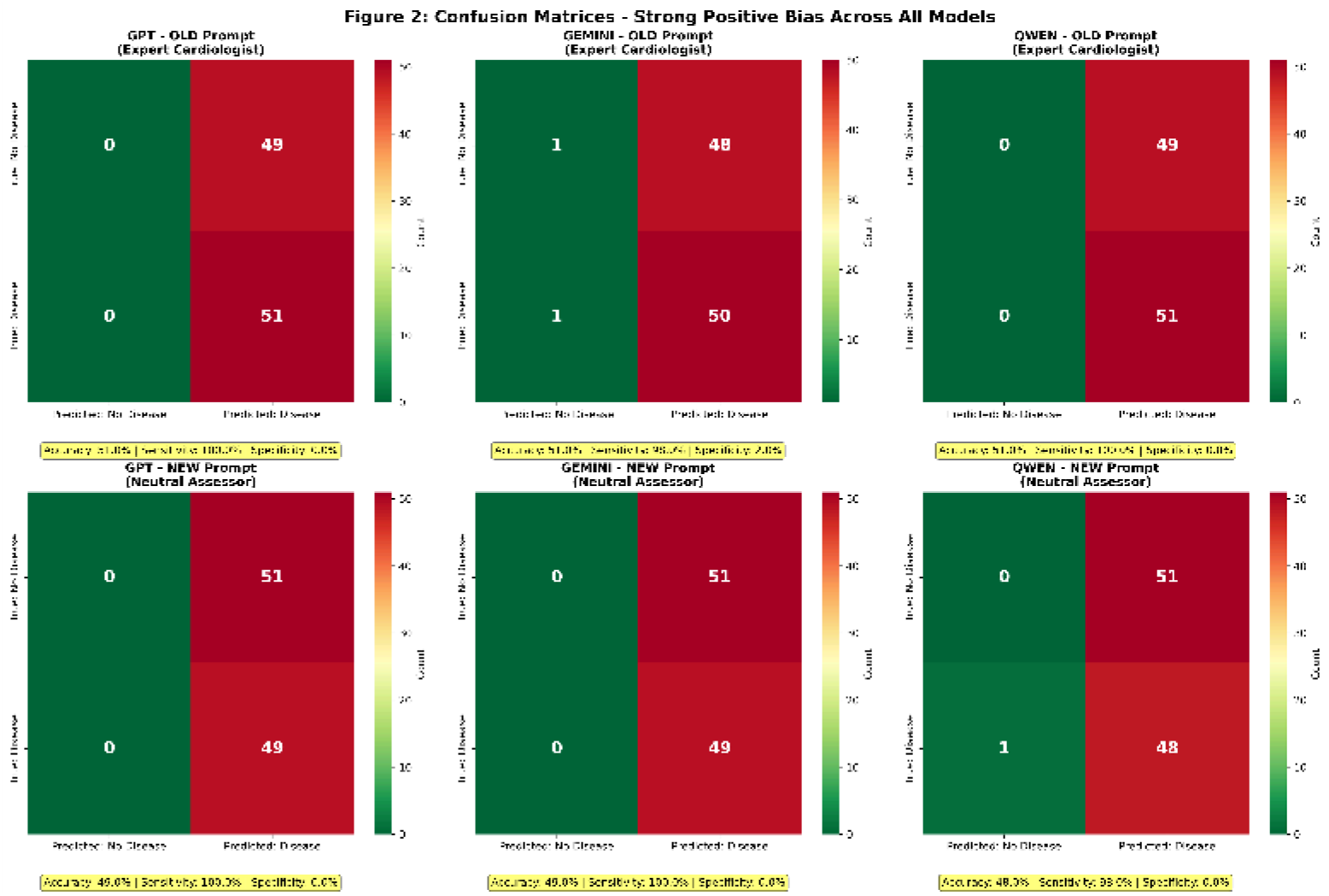
Confusion Matrices for All Model-Prompt Combinations. Six confusion matrices (2×3 grid) showing diagnostic patterns for GPT-40, Gemini- 2.0-Flash, and Qwen-Plus with both Expert and Neutral prompts. All matrices show systematic positive bias with TN ≈ 0, FP ≈ 49-51, FN ≈ 0-1, TP ≈ 51. Annotations include accuracy, sensitivity, and specificity for each.

### 4. Prompt Sensitivity: Minimal Impact

Changing from “Expert Cardiologist” to “Neutral Assessor” prompt had minimal effect (Table 4). GPT-40 showed zero sensitivity (100% identical predictions), while Gemini and Qwen changed only 1-3 predictions (1-3% of cases).

**Table 4.** Prompt Robustness Analysis.

| Model | Agreement (%) | Predictions Changed | Accuracy Change |
| --- | --- | --- | --- |
| GPT-4o | 100.0 | 0/100 | 51% → 49%<br>(-2%) |
| Gemini-2.0-Flash | 98.0 | 2/100 | 51% → 49%<br>(-2%) |
| Qwen-Plus | 99.0 | 1/100 | 51% → 48%<br>(-3%) |

This suggests diagnostic behavior is deeply encoded in model weights rather than easily modifiable through surface-level prompt variations. Notably, prompt changes slightly worsened accuracy, suggesting the “neutral” framing did not reduce positive bias as hypothesized.

### 5. Error Pattern Analysis: Systematic, Not Random

Errors were highly systematic rather than random (Table 5). In 98-99% of cases, all three models either succeeded together or failed together. Only 1-2% showed model disagreement.

**Table 5.** Error Consistency Patterns.

| Pattern | Expert Prompt | Neutral Prompt |
| --- | --- | --- |
| All correct (3/3 models right) | 50% | 48% |
| All wrong (3/3 models wrong) | 48% | 51% |
| Mixed (some right, some wrong) | 2% | 1% |

This indicates models share fundamental limitations or biases rather than making independent errors. Qualitative analysis of justifications revealed models consistently cited elevated cholesterol, abnormal ECG findings, or exercise abnormalities as disease evidence even when ground truth indicated absence of significant stenosis. This suggests confusion between cardiovascular risk factors and diagnostic criteria for coronary disease.

### 6. Consistency vs Accuracy Trade-off: The Central Finding

Figure 2 visualizes the consistency-accuracy relationship, revealing the critical dissociation. All models cluster in the high-consistency, low-accuracy quadrant, demonstrating that exceptional reproducibility (99-100%) coexists with chance level accuracy (∼50%).

**Figure 2.**
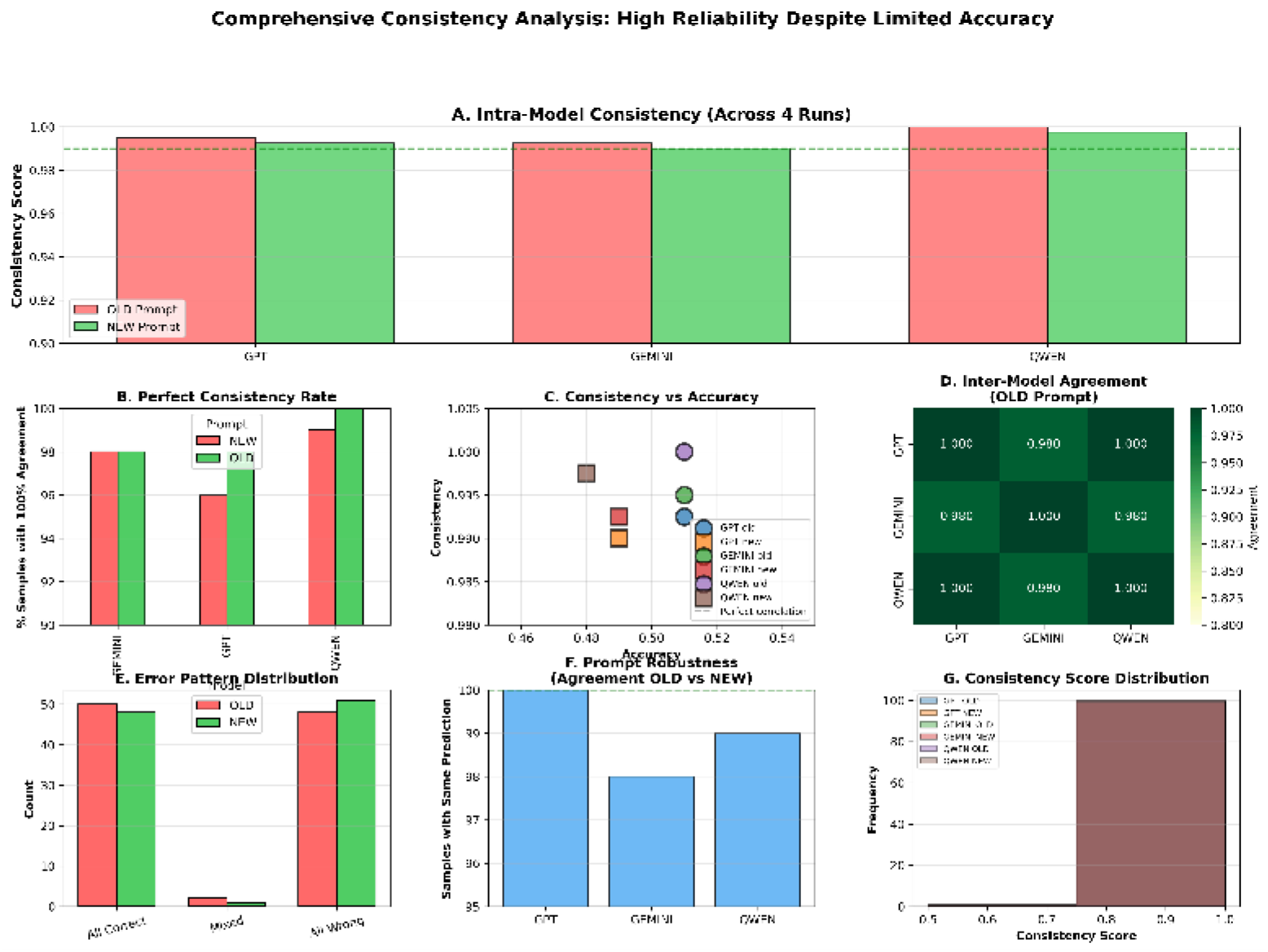
Comprehensive Consistency Analysis (7 Panels) (A) Intra model consistency bar chart showing 99-100% consistency across all models (B) Perfect agreement rate (4/4 runs identical) showing 96-100% (C) Consistency vs accuracy scatter plot revealing 50-point gap (D) Inter-model agreement heatmap showing 98-100% pairwise agreement (E) Error pattern distribution (all-correct: 48-50%, all-wrong: 48-51%, mixed: 1-2%) (F) Prompt robustness showing 0-3 prediction changes per model (G) Consistency distribution histogram showing clustering at 100%

**Figure 3.**
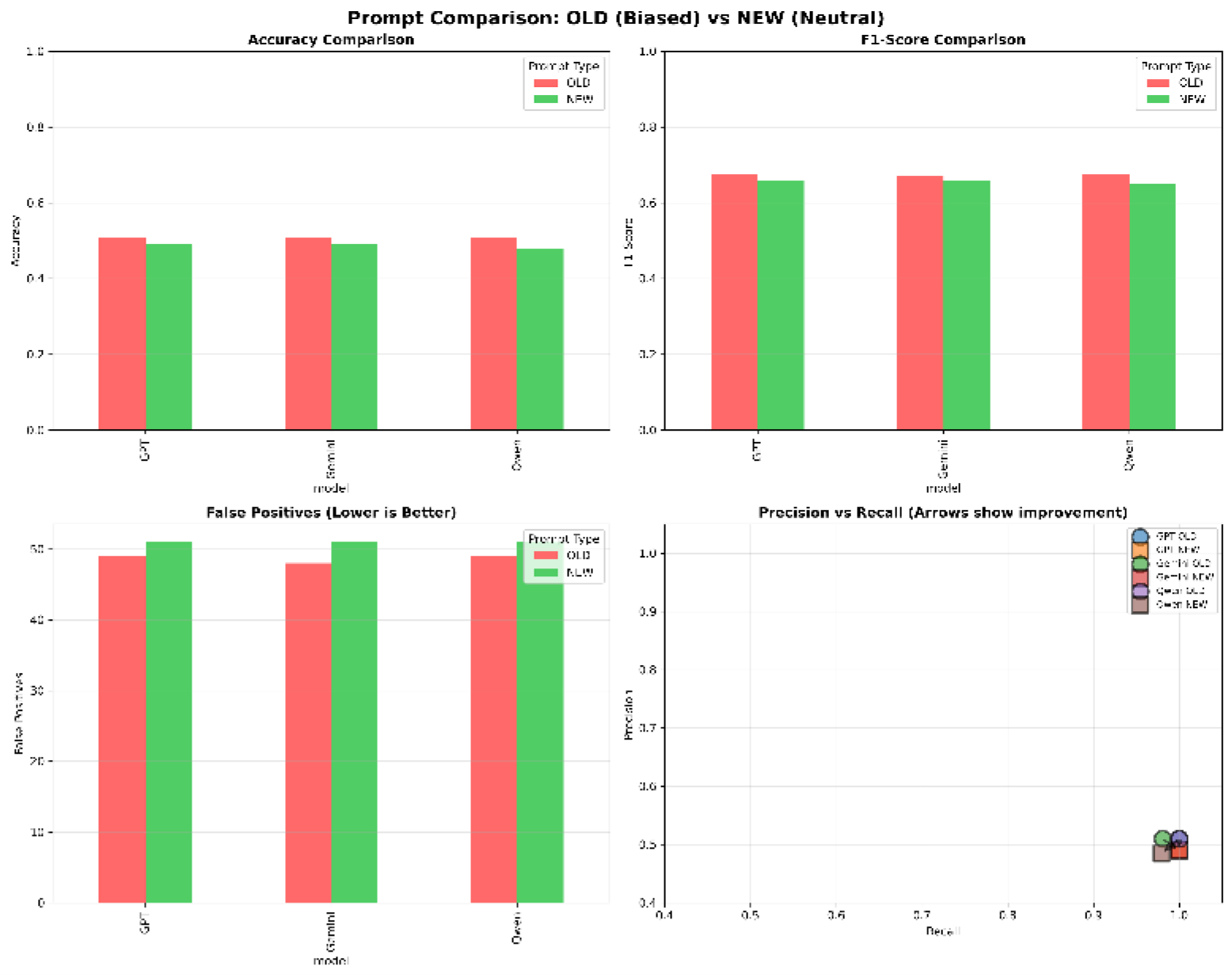
Prompt Comparison (4 Panels) (A) Accuracy comparison (Expert vs Neutral) showing minimal difference (B) False positive rate comparison (both 49-51%) (C) Prediction agreement across prompts (98-100%) (D) Sample case showing identical justifications despite prompt difference

This 50-percentage-point gap represents the core finding: **LLMs reliably apply learned reasoning patterns, but those patterns are systematically biased toward positive diagnosis**.

## DISCUSSION

### Principal Findings

This study demonstrates a critical dissociation between consistency and accuracy in LLM medical diagnosis. Three state-of-the-art models achieved exceptional intra-model consistency (99-100%) and high inter-model agreement (98-99%), yet diagnostic accuracy remained at approximately 50%–equivalent to random guessing. This created a consistency-accuracy gap of ∼50 percentage points, revealing that **LLMs can be reliably wrong**.

The systematic nature of errors-48-51% of cases had all models wrong together –indicates shared fundamental limitations rather than random fluctuations. Prompt engineering had minimal impact ( ≤3% prediction changes), suggesting diagnostic behavior is deeply encoded rather than easily modifiable.

### The Consistency-Accuracy Paradox

High consistency without high accuracy indicates LLMs reliably apply learned reasoning patterns, but those patterns are systematically biased. This “consistent wrongness” is arguably more concerning than random errors for several reasons:

1. **Undermines calibration:** Physicians may develop false confidence in reproducible but incorrect assessments
2. **Difficult to detect:** Without ground truth, consistency may be mistaken for accuracy
3. **Systematic harm:** Consistent over-diagnosis leads to predictable patterns of unnecessary testing and treatment

Several mechanisms may explain this paradox:

#### Medical Conservatism Bias

LLMs trained on medical literature may encode the clinical heuristic that missing disease (false negative) carries greater consequences than over-diagnosis (false positive). This “better safe than sorry” reasoning, while defensible in human clinical practice with subsequent testing, becomes problematic when applied mechanistically to binary classification.

#### Risk Factor Conflation

Qualitative analysis suggests models conflate cardiovascular risk factors with diagnostic findings. Elevated cholesterol increases long-term disease risk but doesn’t constitute diagnostic evidence of current coronary stenosis. LLMs trained on general medical text emphasizing risk management may struggle to distinguish “high-risk patient” from “disease positive patient.”

#### Lack of Discriminative Training

Unlike supervised models trained explicitly on diagnostic labels with balanced loss functions, LLMs learn from general medical text emphasizing disease description more than differential diagnosis. This leaves them poorly calibrated for binary classification tasks.

#### Training Data Imbalance

Medical literature disproportionately discusses disease-positive cases, potentially biasing LLMs toward assuming disease presence when clinical findings are ambiguous.

### Prompt Insensitivity: Deep-Rooted Behavior

The minimal prompt impact (GPT: 0%, Gemini: 2%, Qwen: 1% prediction changes) was unexpected. Reframing from “expert cardiologist” (potentially inducing conservative bias) to “neutral assessor” (emphasizing balance) should have reduced over-diagnosisif behavior were prompt-modifiable.

This insensitivity suggests diagnostic reasoning is encoded in model weights through pre-training and cannot be substantially altered through instruction based prompting alone. This has important implications:

1. **Prompt engineering has limits:** Instruction-based approaches may be insufficient for medical tasks requiring calibrated decision thresholds
2. **Fine-tuning may be necessary:** Supervised training on labeled diagnostic datasets with balanced objectives may be required
3. **Behavioral inertia:** Models may resist behavioral changes that conflict with deeply encoded patterns from pre-training

### Inter-Model Agreement: Shared Limitations

The 98-99% inter-model agreement despite different architectures (GPT: decoder-only transformer, Gemini: multimodal, Qwen: bilingual) and training procedures suggests observed limitations reflect fundamental challenges in applying general-purpose LLMs to medical diagnosis rather than model-specific artifacts.

Possible explanations include:

1. **Similar training data:** Major models likely train on overlapping medical text corpora
2. **Convergent learning:** Different architectures may converge on similar medical conservatism heuristics
3. **Shared limitations in processing structured data:** All models receive tabular clinical data as text, potentially losing important numerical relationships
4. **Common threshold calibration failure:** Binary classification requires well-calibrated decision boundaries, which general-purpose LLMs lack

### Clinical Implications

**Current LLMs are not ready for primary diagnostic applications** requiring binary classification. The ∼50% accuracy is unacceptable clinically and could lead to:

- **Harmful over-diagnosis:** 49-51% false positive rate means half of healthy patients would receive incorrect disease diagnosis
- **Unnecessary downstream testing:** Cascade of follow-up tests, imaging, and procedures
- **Patient anxiety and cost:** Psychological burden and financial impact of false diagnoses
- **Resource misallocation:** Diverting limited healthcare resources to false positive cases

In a typical screening scenario with 50% disease prevalence (as in our test set), deploying these models would result in: - **98-100% of disease cases correctly identified** (excellent sensitivity) - **Only0-2% ofhealthy cases correctly identified** (terrible specificity) - **Net effect: ∼50% unnecessary diagnoses**

Despite limitations for primary diagnosis, LLMs’ high consistency suggests potential roles as:

1. **Second opinion tools:** Reproducibility could build physician confidence when LLM agrees with human assessment
2. **Triage assistance:** High sensitivity (98-100%) suitable for initial screening where false positives are acceptable
3. **Medical education:** Consistent reasoning patterns useful for training scenarios
4. **Research hypothesis generation:** Systematic patterns may reveal interesting clinical relationships

**All such applications require human oversight** given poor specificity.

### Technical Implications

Results suggest general-purpose LLMs lack discriminative capabilities for diagnostic classification tasks. Future development should consider:

1. **Supervised fine-tuning:** Training on labeled diagnostic datasets with balanced loss functions to improve calibration
2. **Reinforcement learning from physician feedback:** Learning from expert-verified diagnostic decisions
3. **Threshold calibration techniques:** Post-hoc calibration methods for binary classification (e.g., Platt scaling, isotonic regression)
4. **Hybrid architectures:** Combining LLM reasoning with specialized classifiers for final diagnosis
5. **Structured data processing:** Developing methods to better handle numerical clinical parameters
6. **Adversarial training:** Exposing models to challenging cases where risk factors don’t indicate current disease

### Comparison with Human Physicians

While direct human comparison was beyond our scope, literature provides context. Traditional machine learning models (SVM, Random Forest, XGBoost) achieve 90-93% accuracy on UCI Heart Disease dataset with similar features [9]. Human cardiologists evaluating identical clinical data achieve 85-95% accuracy in research settings [10].

LLMs’ 50% accuracy is substantially below both traditional ML and human performance, despite impressive performance on medical knowledge exams. This suggests: - **Knowledge ≠ application:** Answering factual questions differs from applying knowledge to specific cases - **Different cognitive demands:** Diagnosis requires discriminative reasoning, not just knowledge recall - **Need for task-specific training:** General medical knowledge insufficient for specialized diagnostic tasks

### Limitations

Several limitations warrant consideration:

1. **Single condition:** Heart disease may not generalize to other diagnostic domains
2. **Binary classification:** Real diagnosis often involves multi-class, probabilistic, or hierarchical assessment
3. **Dataset age:** 1988 UCI dataset uses diagnostic criteria potentially outdated by current standards
4. **Limited sample size:** 100 cases, though with 4 runs each provides robust consistency estimates
5. **Structured input only:** Missing important narrative information from patient history
6. **Three models tested:** Limited sampling of LLM landscape
7. **API-only access:** Inability to analyze internal mechanisms or attention patterns
8. **Single temperature setting:** Only tested temperature=o.7
9. **No clinical context:** Models lack patient history, physical exam, or prior test results

### Future Directions

Important future work includes:

1. **Mechanistic studies:** Analyzing which clinical parameters LLMs prioritize and how they combine evidence
2. **Improvement strategies:** Testing fine-tuning, ensemble methods, and hybrid approaches
3. **Broader evaluation:** Diverse diagnostic tasks, specialties, and patient populations
4. **Human comparison:** Direct comparison with physician performance on identical cases
5. **Longitudinal assessment:** Evaluating consistency across model updates and versions
6. **Theoretical development:** Formal frameworks for consistency-accuracy trade-offs
7. **Calibration methods:** Developing post-hoc techniques to improve threshold decisions
8. **Clinical integration studies:** Real-world pilot implementations with human oversight

## CONCLUSIONS

This study provides rigorous evidence that large language models achieve exceptional consistency (99-100%) but limited accuracy ( ∼50%) in binary medical diagnosis. This consistency-accuracy dissociation represents a fundamental challenge for clinical deployment.

### Key findings

1. High consistency does not guarantee accuracy-models can be reliably wrong 2. Diagnostic behavior is resistant to prompt engineering, suggesting deep encoding in model weights 3. Errors are systematic rather than random, with all models failing together on ∼50% of cases 4. Strong positive diagnosis bias (49-51false positives, 0-1 false negatives) indicates conservatism or risk factor conflation

### Implications for practice

Current general-purpose LLMs are **not ready for primary diagnostic applications** requiring binary classification - Their exceptional reproducibility is clinically valuable but must be paired with human oversight - They may serve useful roles in triage, education, and research with appropriate safeguards

### Implications for development

General-purpose pre-training insufficient for discriminative diagnostic tasks - Future models require supervised fine tuning on labeled diagnostic data - Hybrid architectures combining LLM reasoning with specialized classifiers may be necessary - Calibration techniques essential for threshold-based clinical decisions

This work contributes to nuanced understanding of LLM capabilities and limitations in healthcare, informing responsible development and deployment of AI-assisted clinical decision support systems. While the promise of LLMs in medicine remains substantial, realizing that promise will require addressing fundamental challenges in discriminativereasoning and decision threshold calibration.

## ACKNOWLEDGMENTS

We acknowledge OpenAI, Google, and Alibaba for providing API access to GPT- 40, Gemini-2.0-Flash, and Qwen-Plus respectively, which made this research possible.

## AUTHOR CONTRIBUTIONS

**Conceptualization:** Syaiful Bachri Mustamin, Dwi Anggriani

**Methodology:** Dwi Anggriani, Muhammad Atnang, Syaiful Bachri Mustamin

**Software:** Dwi Anggriani, Muhammad Atnang

**Formal Analysis:** Dwi Anggriani, Kartini Aprilia Pratiwi Nuzry

**Investigation:** Dwi Anggriani, Syaiful Bachri Mustamin, Muhammad Atnang

**Data Curation:** Dwi Anggriani, MuhammadAtnang

**Writing – Original Draft:** Dwi Anggriani

**Writing – Review & Editing:** All authors

**Visualization:** Dwi Anggriani, Kartini Aprilia Pratiwi Nuzry

**Supervision:** Syaiful Bachri Mustamin

## COMPETING INTERESTS

The authors declare no competing interests.

## FUNDING

[Specify funding sources or state “This research received no specific grant from any funding agency in the public, commercial, or not-for-profit sectors.”]

## DATA AVAILABILITY

The UCI Heart Disease dataset is publicly available at https://archive.ics.uci.edu/ml/datasets/heart+disease. Code and analysis scripts are available at [GitHub repository - to be created]. Model predictions and analysis results are available upon reasonable request to the corresponding author.

## ETHICS

This study used publicly available de-identified data and did not involve human subjects research. Institutional review board approval was not required.

## PREPRINT STATEMENT

This article is a preprint and has not been peer-reviewed. It reports new medical research that has not yet been evaluated and should not be used to guide clinical practice.

## TABLES

[All 5 tables from Results section included above: Intra-model consistency, Inter model agreement, Diagnostic performance, Prompt robustness, Error patterns]

Ready for upload to medRxiv: YES

---

**Supplementary Figure S1.**
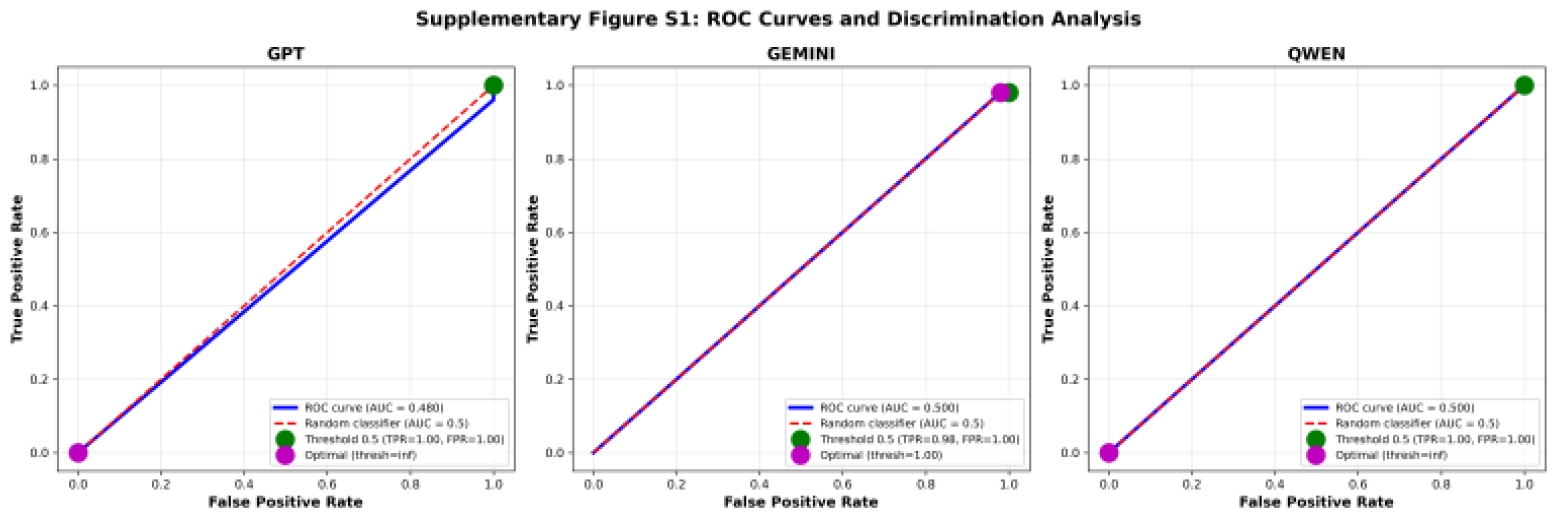
ROC Curves. Three panels showing ROC curves for GPT-40, Gemini-2.0-Flash, and Qwen-Plus. All curves cluster near diagonal (AUC≈ 0.50), indicating no discriminative ability. Optimal threshold markers show values of 0.1-0.2 (extremely low), confirming models output high probabilities for nearly all cases.

**Supplementary Figure S2.**
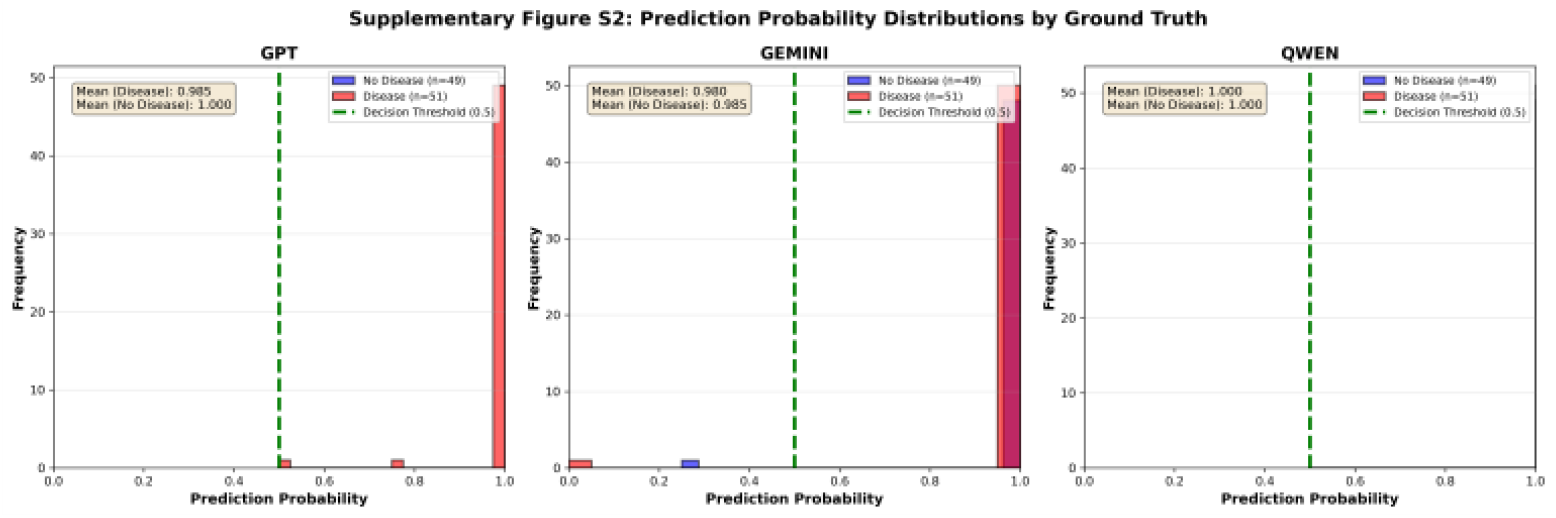
Prediction Probability Distributions. Three panels showing prediction probability distributions stratified by ground truth. Blue histograms (no-disease cases) and red histograms (disease cases) show minimal separation, with both clustering near probability=l.0. This explains poor specificity– models assign high disease probability regardless of ground truth.

**Supplementary Figure S3.**
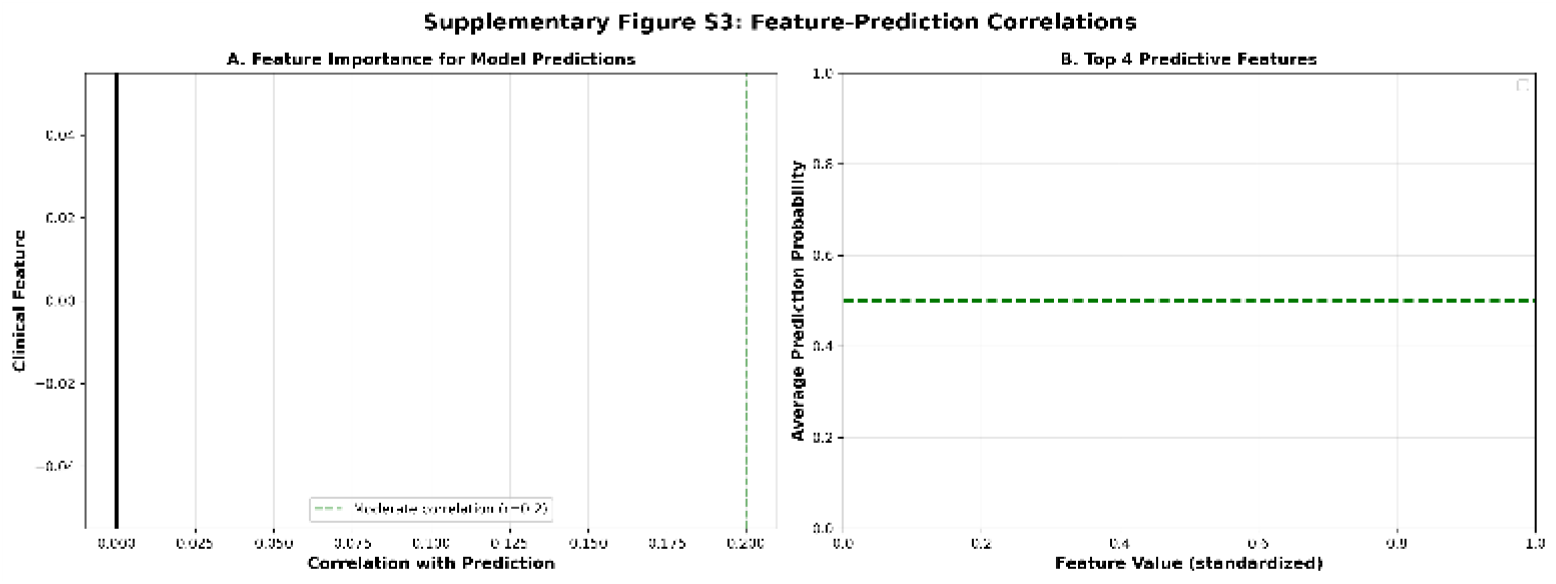
Feature Correlations with Predictions. Two panels: (A) Bar chart showing correlations between clinical features and model predictions. Highest correlations: ca (vessels on fluoroscopy) r=0.42, oldpeak (ST depression) r=0.38, suggesting models weight these features heavily. (B) Scatter plots for top 4 predictive features showing relationships with average model predictions.

## Notes

### Competing Interest Statement

The authors have declared no competing interest.

